# Infectious disease dynamics and restrictions on social gathering size

**DOI:** 10.1101/2022.01.07.21268585

**Authors:** Christopher B Boyer, Eva Rumpler, Stephen M Kissler, Marc Lipsitch

**Author notes:** C. B. and E R. are co-first authors.

## Abstract

Social gatherings can be an important locus of transmission for many pathogens including SARS-CoV-2. During an outbreak, restricting the size of these gatherings is one of several non-pharmaceutical interventions available to policy-makers to reduce transmission. Often these restrictions take the form of prohibitions on gatherings above a certain size. While it is generally agreed that such restrictions reduce contacts, the specific size threshold separating “allowed” from “prohibited” gatherings often does not have a clear scientific basis, which leads to dramatic differences in guidance across location and time. Building on the observation that gathering size distributions are often heavy-tailed, we develop a theoretical model of transmission during gatherings and their contribution to general disease dynamics. We find that a key, but often overlooked, determinant of the optimal threshold is the distribution of gathering sizes. Using data on pre-pandemic contact patterns from several sources as well as empirical estimates of transmission parameters for SARS-CoV-2, we apply our model to better understand the relationship between restriction threshold and reduction in cases. We find that, under reasonable transmission parameter ranges, restrictions may have to be set quite low to have any demonstrable effect on cases due to relative frequency of smaller gatherings. We compare our conceptual model with observed changes in reported contacts during lockdown in March of 2020.

## 1 Introduction

Social gatherings in which people meet and interact provide a conducive environment for the spread of pathogens. During an outbreak, restricting the size of such gatherings is one of several nonpharmaceutical interventions (NPIs) available to policymakers. An advantage of these restrictions is that they are simple to articulate and easy for the public to understand and, in some circumstances, for authorities to enforce. Indeed, during the COVID-19 outbreak, gathering size restrictions were among the most commonly used NPIs globally (Hale et al., 2021). Some have claimed that these restrictions were among the most effective at reducing transmission (Brauner et al., 2021; Haug et al., 2020; Li et al., 2021; Sharma et al., 2021); however, given the rapid and often haphazard nature of their rollout and the methodological challenges of proper indentification, estimates of the causal effects of specific NPIs may be severely biased (Haber et al., 2021).

Theory and intuition suggest that, when properly followed, gathering size restrictions should lower transmission by limiting the number of contacts between individuals and thus reducing the opportunity for the pathogen to spread. Yet, it’s often unclear precisely how low restrictions should be set to achieve a certain disease control target, be it a stable number of cases or the elimination of the pathogen from the population. Indeed, evidence suggests, policymakers have taken a variety of approaches in practice to set gathering size thresholds, which may reflect different goals or local disease dynamics, but also might reflect ambiguity in the optimal strategy. As a case in point, in the UK the government first banned gatherings above 500 in March 2020 before initiating a lockdown on March 23. Then after restrictions eased in the late summer a ban on gatherings above 30 was declared, but then this was famously revised down to the “rule of six” in September to prevent gatherings with more than six people. In this paper, we use epidemiological theory to better understand the relationship between gathering size and general disease dynamics. We also attempt to enumerate the necessary elements to quantify or predict the impact of a given threshold on the incidence of new cases.

We emphasize restrictions on gathering sizes for several reasons. Firstly, we note that a significant proportion of the superspreading events in the literature, including some of the most spectacular accounts, have occurred during social gatherings. Given the outsized role these events seem to play at the start of outbreaks, some have hypothesized that control and/or suppression of an emerging pathogen could largely be achieved via targeted reduction in mass gatherings. Secondly, several retrospective reports comparing confirmed COVID-19 cases and test-negative controls (*COVID-19: Note by the Chief Medical Officer, Chief Nursing Officer and National Clinical Director*, 2020; Fisher et al., 2020) have found an association between attending family and friends gatherings and infection with SARS-CoV-2, suggesting that gatherings may be important source of new cases. Thirdly, social gathering restrictions seem to be among the first and most frequent measures to be implemented, which perhaps can be explained by a perception that social gatherings have less social value than other gatherings that occur in venues such a schools and hospitals. Finally, while both the United States and European Centers for Disease Control recommended limiting the size and duration of gatherings (Center for Disease Control and Prevention (CDC), 2020; European Center for Disease Control (ECDC), 2020), the specific timing of when to implement restrictions and the numeric threshold separating “allowable” from “prohibited” gatherings generally did not have a clear scientific basis. This lead to dramatic differences in guidance across location and time. For instance, while most countries implemented limitations in the spring of 2020, the intensity and duration of these restrictions varied extensively from country to country (Roser et al., 2020) with maximum gathering sizes permitted ranging from 2 to 5000 and subject to frequent and somewhat erratic changes as the epidemic progressed.

## 2 Theory

As in the standard compartmental model, we consider the epidemic spread of a pathogen in a population which can be divided into three disjoint sets of individuals: susceptible and not yet infected (*S*), infected and infectious (*I*), or recovered, no longer infectious, and immune (*R*). As time passes, individuals in the population come into contact with one another and the pathogen spreads through contacts between susceptible and infectious individuals. Gathering size restrictions limit the number of contacts that individuals have, but apply only to a subset of contacts that occur during social gatherings. Therefore, we categorize all contacts between individuals as either occurring during “gatherings”, i.e. non-household settings that are presumably affected by gathering size restrictions, or at home or other settings not affected by gathering size restrictions, and we focus on the former as the source of the contribution of gatherings to disease dynamics.

At each time point, individuals attend *M* gatherings of size *K*, where *K* is a random variable defined by some distribution *f*(*k*). To simplify matters here we include the possibility that an individual does not attend a gathering in *f*(*k*) through defining it as a gathering size of 1 so that the same distribution applies to everyone. We define *X* to be the number of incident cases. We assume for now that policies target the expected number of new infections that can be attributed to gatherings, *E*(*X*), which, using the law of total probability, can be written as:

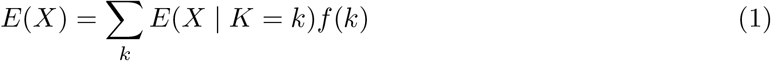

where *E*(*X* | *K* = *k*) is the expected incidence of new infections at a given gathering size of *K* = *k*. This expression suggests that we could write the expected rate under an idealized restriction, that is a restriction that is strictly enforced such that no one attends a gathering with *k* > *k*_*max*_ as:

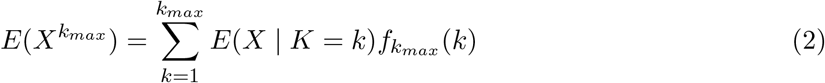

where the sum is now over the restricted range of gathering sizes and 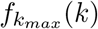 is the distribution of gathering sizes after the restriction has been applied recognizing that it could differ from simply a truncated *f*(*k*) as people may respond to the restriction in different ways. Therefore, in order to estimate the potential impact of a gathering size restriction, we need two essential inputs: (1) the distribution of gathering sizes and (2) the relationship between gathering size and expected number of infections. Then, given a range of *k*_*max*_ values, policymakers could ideally target a specific reduction in new cases *X*^∗^, and select *k*^∗^ = max(*k*_*max*_) such that *X* < *X*^∗^, perhaps weighing them against the cost of imposing the restriction.

Starting with the second input, as we show in section A.1.1 of the Appendix, the expected number of incident cases *X* that occur at a gathering of size *K* = *k* is :

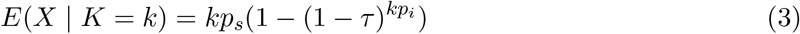

where *τ* is the probability of transmission given contact, and *p*_*s*_ and *p*_*i*_ are the population proportions of susceptible and infectious individuals respectively. We assume susceptible, infectious, and recovered individuals attend gatherings at rates roughly equivalent to their population proportions and that everyone who attends a gathering comes into contact with all other attendees. Figure 1a plots this expression for example values of *τ, p*_*i*_, and *p*_*s*_. As intuition might suggest, it shows that, for a single gathering, larger gatherings produce more secondary infections than smaller gatherings and that this relationship is nonlinear as larger gatherings both increase the number of potential contacts and as well as the expected number of infectious individuals in attendance. Indeed, as shown in section A.1.2 of the Appendix, for small values of *τ* and *p*_*i*_ that are typical of an infectious disease outbreak, i.e. |*τkp*_*i*_| ≪ 1, we can use a Binomial approximation to simplify this to:

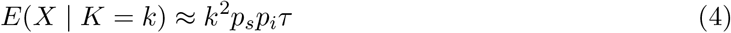

which is quadratic in the size of the gathering.

**Figure 1:**
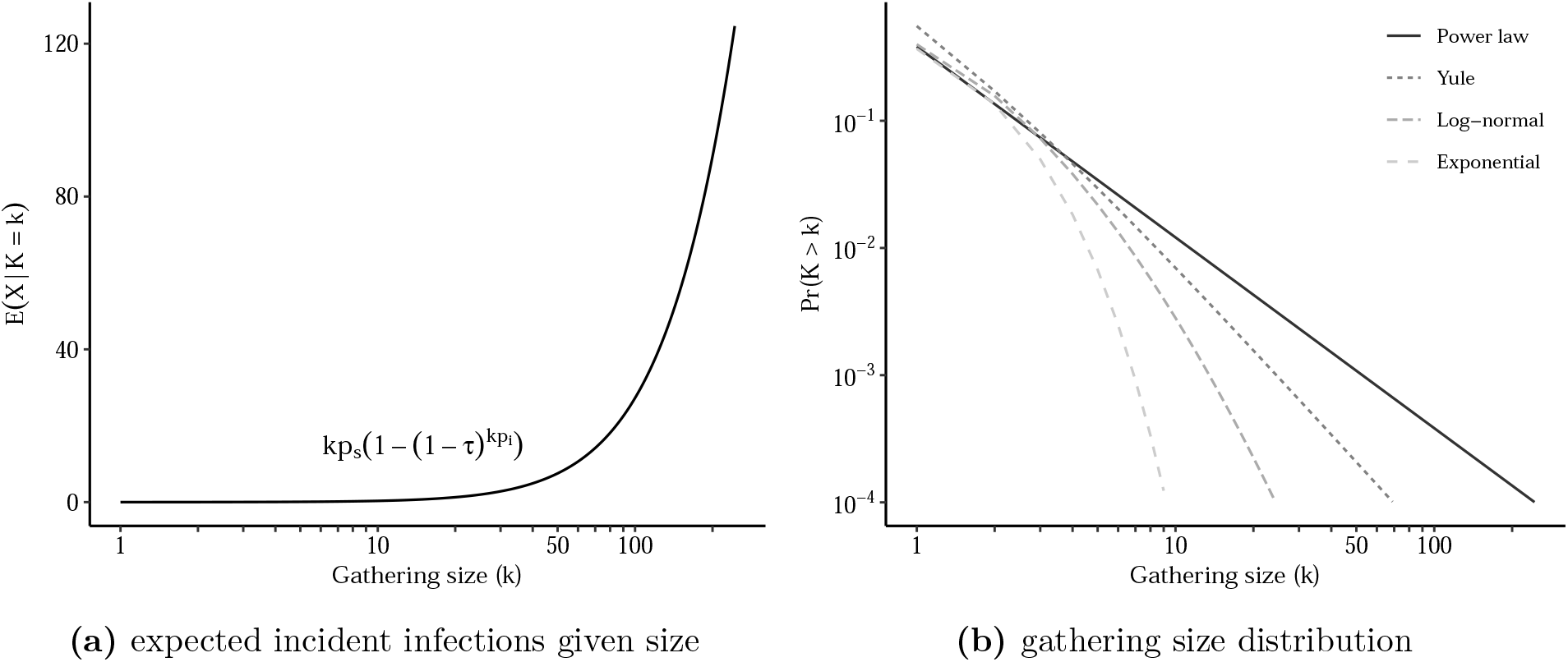
Plots of necessary components of effect of gathering size restriction. In panel (a), we fix transmission parameters to following values *p*_*i*_ = 0.01, *p*_*s*_ = 0.99, *τ* = 0.08 and assume power law behavior starts at *k*_*min*_ = 1.

As for the other input, the distribution of gathering sizes, empirical studies suggest that human contact distributions may be subexponential, or even scale-free or heavy-tailed, with considerable probability mass in the extreme tail of the distribution (Bansal et al., 2007; Barabási & Albert, 1999; May & Lloyd, n.d.; Pastor-Satorras & Vespignani, 2001). This observation applies equally to distributions of gathering size, i.e. *f*(*k*), as most gatherings are small, but gatherings of tens or hundreds of thousands of individuals are possible. Several generative models of human social interaction have been proposed to explain this phenomena based on random walks (Kelker, 1973) or attracting sites (Barabási & Albert, 1999). Figure 1b shows a few common examples of heavy-tailed distributions. In the extreme case, the limit or asymptotic behavior of these distributions can be characterized by a discrete power law of the form

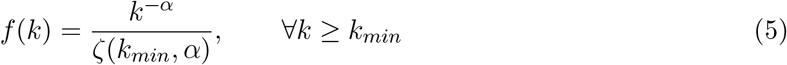

where 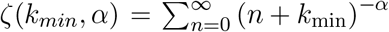 is generalized zeta function and *k*_*min*_ is the threshold for power-law behavior. This has important implications as moments of power law distributions may not be finite under some parameterizations as the extreme mass in the tail leads to infinite sums or integrals. For instance, it is well known that the number of finite moments of power-law distributions is determined by the value of *α*, when *α* < 3 the distribution has finite mean but infinite variance and when *α* < 2 the distribution has no finite moments. Many observed phenomena exhibit power-law behavior with 2 ≤ *α* ≤ 3 (Clauset et al., 2009).

Assuming that, in the range of *k*_*max*_ restrictions considered, a power law is a good approximation for the distribution of gathering sizes, and combining this with the results in equation 3 and 4, the expected rate of new infections under restriction simplifies to:

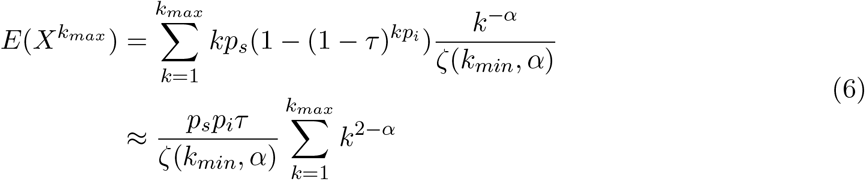

Viewing the 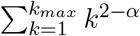 as a weighted sum denoting the contribution of gatherings of size between 1 and *k*_*max*_ to the rate of new infections yields the following insight: when *α* < 2 the contributions are increasing suggesting that larger gatherings contribute more to the rate of new infections than smaller gatherings and by extension there are diminishing returns to imposing lower restrictions; while, on the other hand, when *α* ≥ 2 contributions are flat or decreasing suggesting that smaller gatherings contribute more to the rate of new infections than larger gatherings and by extension there are increasing returns to imposing lower restrictions.

In Figure 2 we plot an example of the relative rate of incident cases under a restriction which prohibits gatherings above size *k*_*max*_ for power law distributions of gathering size with different *α* values. Here, we see that when *α* is 2 or below, restrictions of larger gatherings quickly leads to a large reduction in cases; however, as *α* increases vastly more stringent restrictions are required to achieve meaningful reductions. This suggests that the empirical distribution of gathering sizes and the tail-behavior specifically, i.e. the frequency of very large gatherings relative to small ones, are important parameters in determining the optimal threshold for gathering size restrictions. As shown in section A.2.1 and A.2.2, these results are robust to variations in *τ* and *p*_*i*_ and *p*_*r*_, respectively. In A.2.3 we relax the assumption that the distribution of gatherings follows a power law, and instead consider a log-normal distribution.

**Figure 2:**
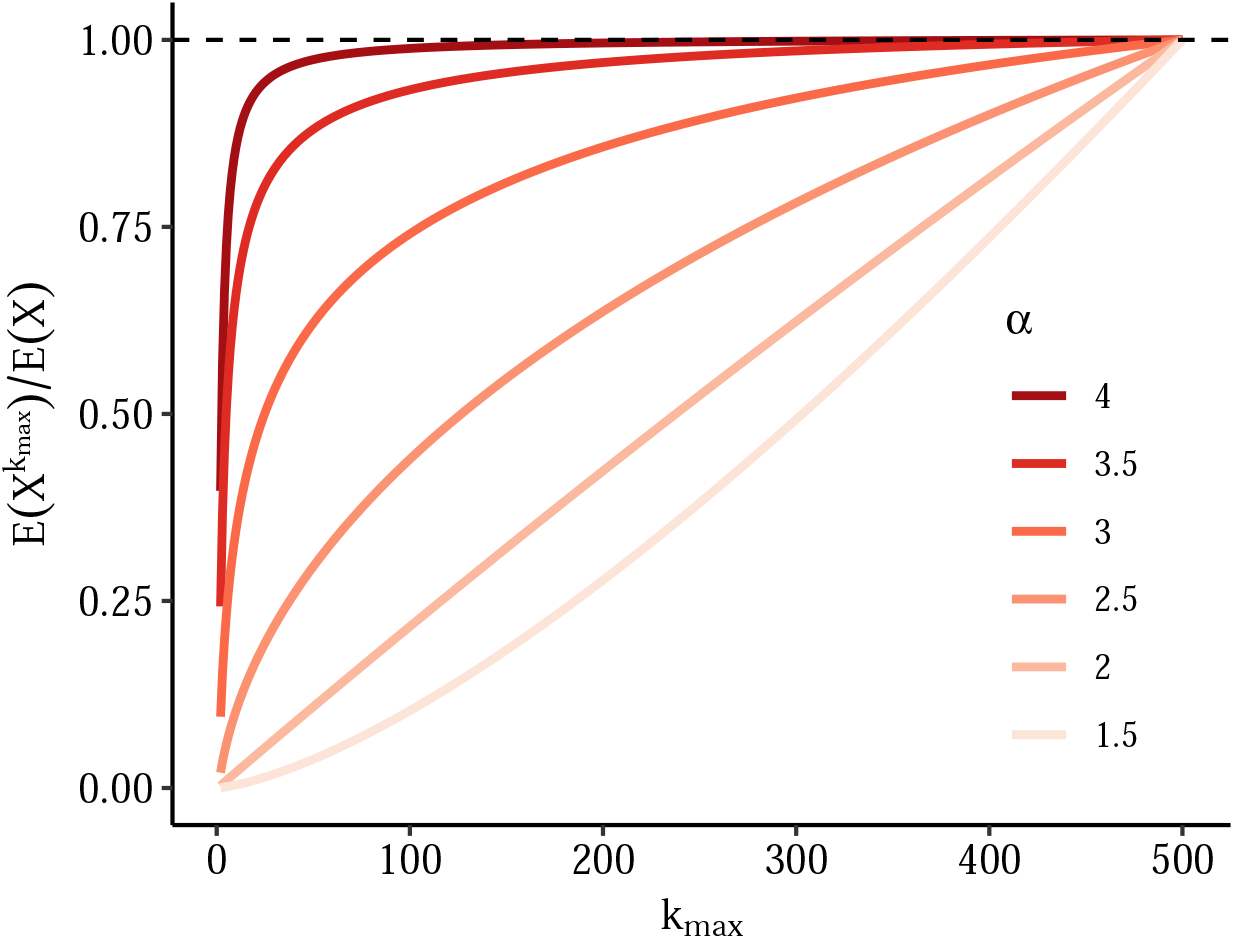
Relative rate of incident cases under restriction which prohibits gatherings above size *k*_*max*_ for different power law distributions. Here we fix transmission parameters to following values *p*_*i*_ = 0.01, *p*_*s*_ = 0.99, *τ* = 0.08 and assume power law behavior starts at *k*_*min*_ = 1. We truncate the power law above gatherings of size 500 both to make the sum tractable and given that gathering sizes must at minimum be less than population size. The figure shows the relative rate of incident cases calculated using equation 6 and comparing restrictions with *k*_*max*_- level thresholds to unrestricted rate (e.g. a value of 0.5 implies a 50% fewer per capita incident cases at time *t* relative to unrestricted rate).

## 3 Application

In the previous section, we showed the distribution of gathering sizes, and the tail-behavior more specifically, is an important determinant of the degree to which smaller or larger gatherings contribute to epidemic dynamics. In this section, we use observational data on the size of human gatherings from multiple sources to estimate the empirical power law behavior of gathering size distributions. We use data collected both during “normal” times and during the COVID-19 pandemic as a reference for *f*(*k*) an 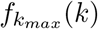 respectively. We estimate the effect of gathering size restrictions during the COVID-19 pandemic using the results from the previous section and empirical estimates of transmission parameters.

Our data on gathering size distributions in the pre-pandemic period are from two primary sources: the BBC Pandemic study (Klepac et al., 2018) and the Copenhagen Networks Study (CNS) of university sources (Sekara et al., 2016). Both are described in more detail elsewhere. Briefly, the BBC Pandemic study is a citizen science project in which UK citizens self-reported daily contacts using a mobile app in 2018. We extracted the number of contacts made in a day by setting (home, work/school or other) for over 38,000 participants (Kucharski et al., 2020) and then divide by the gathering size to account for the over selection of large gatherings. In the Copenhagen Networks Study, the movement and contacts among approximately 1000 university students were intensively tracked and measured via Bluetooth, telecommunication networks, online social media contacts and geolocation over a 5 month period in 2014. In the supplement to the original study the authors report the distribution of 23,231 gatherings observed during the study period. A gathering was defined as groups of individuals in close physical proximity that persists for at least 20 minutes. We extracted the raw data for the probability of observing gatherings of different sizes (Supplementary Figure S9a) using `WebPlotDigitizer`, an online tool that allows the extraction of numerical data from graphs (Rohatgi, 2020).

Table 1 provides the descriptive statistics for the empirical distributions of gathering sizes extracted from both sources. In all cases except for household sizes the variance is much greater than the mean which is indicative of overdispersion or a “heavy tail”. The maximum gathering sizes outside the household were between 200 and 315. The 90th and 99th empirical quantiles similarly suggest extreme skewness.

**Table 1:** Descriptive statistics for empirical distributions of gathering size.

| Data source | N | mean | variance | q90 | q99 | min | max |
| --- | --- | --- | --- | --- | --- | --- | --- |
| BBC Pandemic |  |  |  |  |  |  |  |
| Home | 16,524 | 2.3 | 1.4 | 4 | 6 | 1 | 10 |
| Work / school | 19,044 | 2.0 | 10.0 | 4 | 15 | 1 | 235 |
| Other | 19,459 | 2.0 | 5.5 | 4 | 12 | 1 | 201 |
| Total | 55,026 | 2.1 | 5.8 | 4 | 12 | 1 | 235 |
| Copenhagen Networks Study | 58,227 | 4.9 | 103.1 | 7 | 46 | 2 | 315 |

Figure 3 plots the full distribution of gathering sizes from both sources using a log-log scale. In all contexts the majority of individuals have very few contacts. For work/school and social gatherings, a very long tail of individuals have very large number of encounters (up to 234 daily contacts at work). We plot both the empirical mass function and the complementary cumulative distribution function (CCDF), also often referred to as Zipf plot, noting that the second is generally preferred for distinguishing power-law type behavior. Typically, a CCDF plot from a power law should be linear on a log-log scale. Here we see that most contexts exhibit approximately linear behavior over significant range; however at the extreme right there may be some nonlinearity which may suggest the presence of an upper bound (for instance the gathering size cannot exceed population size). Interestingly, the distributions of gathering size reported in the Copenhagen Networks Study (CNS) and number of contacts reported in the BBC Pandemic study are similar in range and shape, despite having been measured using completely different methodologies at different times and locations.

**Figure 3:**
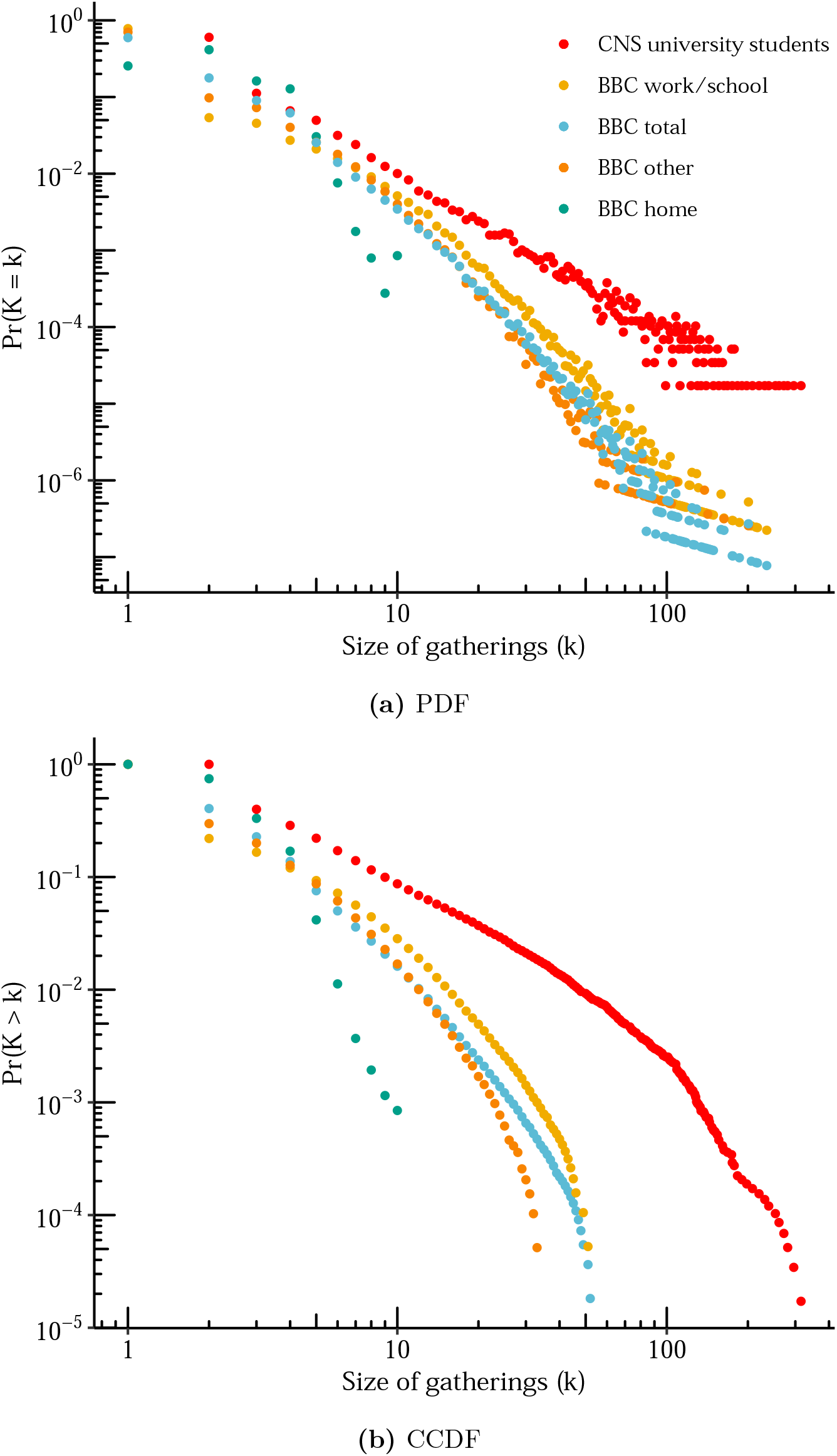
Distribution of gathering sizes from the CNS and the BBC Pandemic study by setting. Empirical distribution of gathering size from the CNS as well as the BBC Pandemic study by setting (home, work/school, other and total). Panel (a) is a log-log plot of the empirical probability that each size is observed. Panel (b) is the empirical complementary cumulative distribution function, i.e. the probability of observing size greater than or equal to *k*, and is often preferred for understanding tail behavior.

Next, we find the best fitting power law for the observed distributions using maximum likelihood. Using the `poweRlaw` (Gillespie, 2014) package in R, we estimate *α* as well as *k*_*min*_ representing the size beyond which the distribution exhibits power law behavior. For the latter, we use the approach of Clauset et al. (Clauset et al., 2009) and estimate it by finding the value which minimizes the Kolmogorov-Smirnov statistic. We estimate the standard errors for both using the bootstrap.

Figure 4 shows the resulting power law fits for each of the data sources. The *α* values estimated range from 2.28 to 6.94, with all settings other than households between 2 and 4. The estimate for the Copenhagen Network study in particular is consistent with infinite second moments (i.e. infinite variance). However, visual inspection suggests that a single power law might not fit well in the extreme tail, with most settings exhibiting considerably lower observed frequencies than suggested by the best-fitting power law. This may be partially due to low cell counts or sampling variability in these extreme quantiles, or as discussed previously may be reflective of the fact that the true distribution is truncated with an upper bound on gathering size.

**Figure 4:**
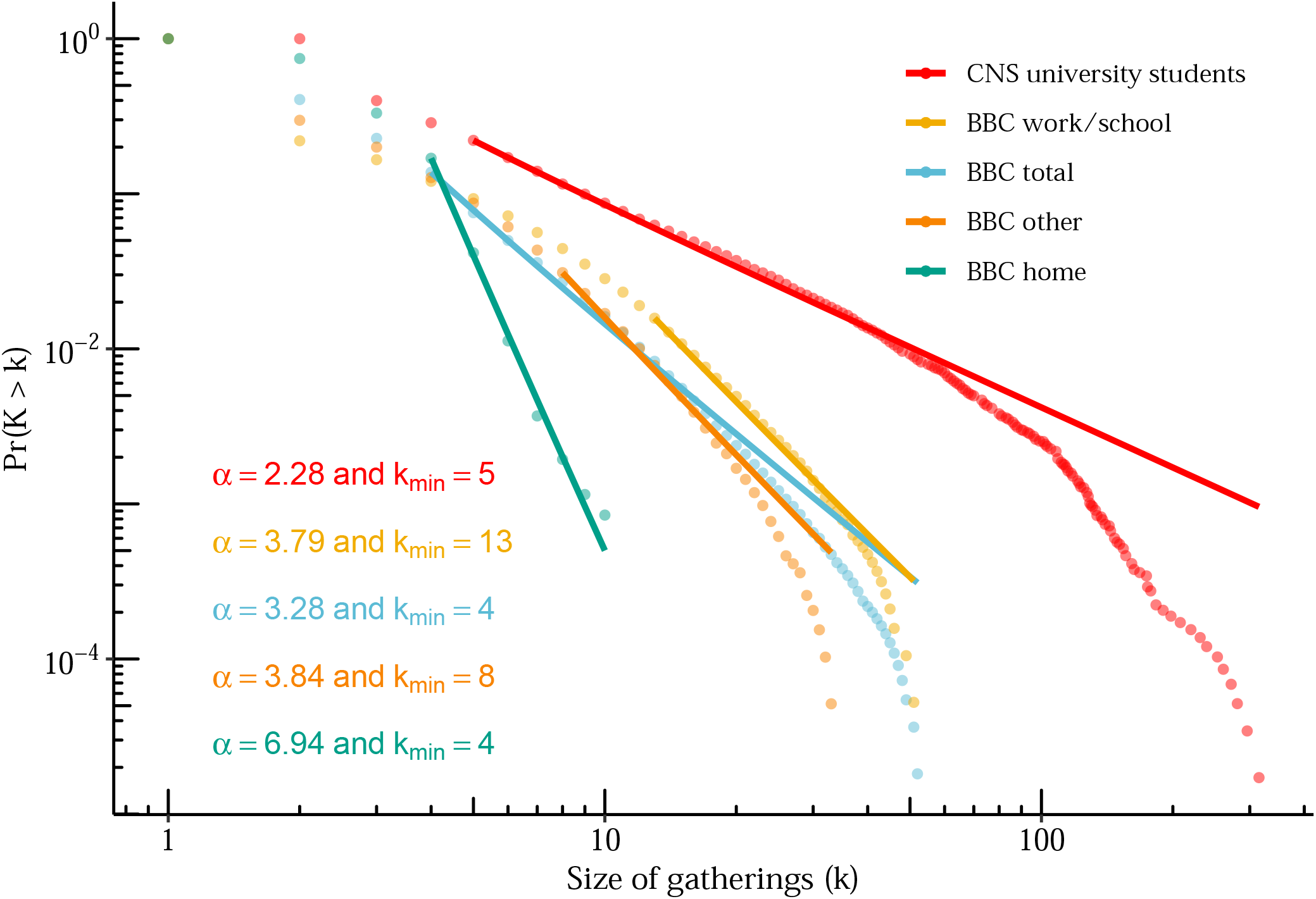
Estimates of power law parameters for the Copenhagen Networks Study (CNS) and the BBC Pandemic study by setting. Plot is complementary cumulative distribution function versus gathering size with lines showing fitted power law distribution. Estimates for *α* and *k*_*min*_ obtained using maximum likelihood for discrete power law using the `poweRlaw` package in R.

Next, we attempt to estimate the effect of a hypothetical gathering size restriction by replicating the analysis shown in Figure 2 but substituting the empirical gathering size distributions. This would represent an idealized intervention in which everyone followed the restriction by not attending a gathering over the threshold, but their other gathering-seeking behavior is otherwise unaffected. Figure 5 shows the results for the distributions in each of the data sources. Here we see that, to achieve reduction in cases of 50% or more, restrictions must be set below 30 in most settings. Compared with the results in Figure 2, however, we see that the empirical distributions suggest a larger impact of restrictions on medium to large size gatherings, likely because the empirical distributions have slightly less mass in the extreme tail than would be suggested by a true power law. As shown in A.2.2, these results remain mostly unchanged when allowing values of *p*_*i*_ and *p*_*r*_ to vary.

**Figure 5:**
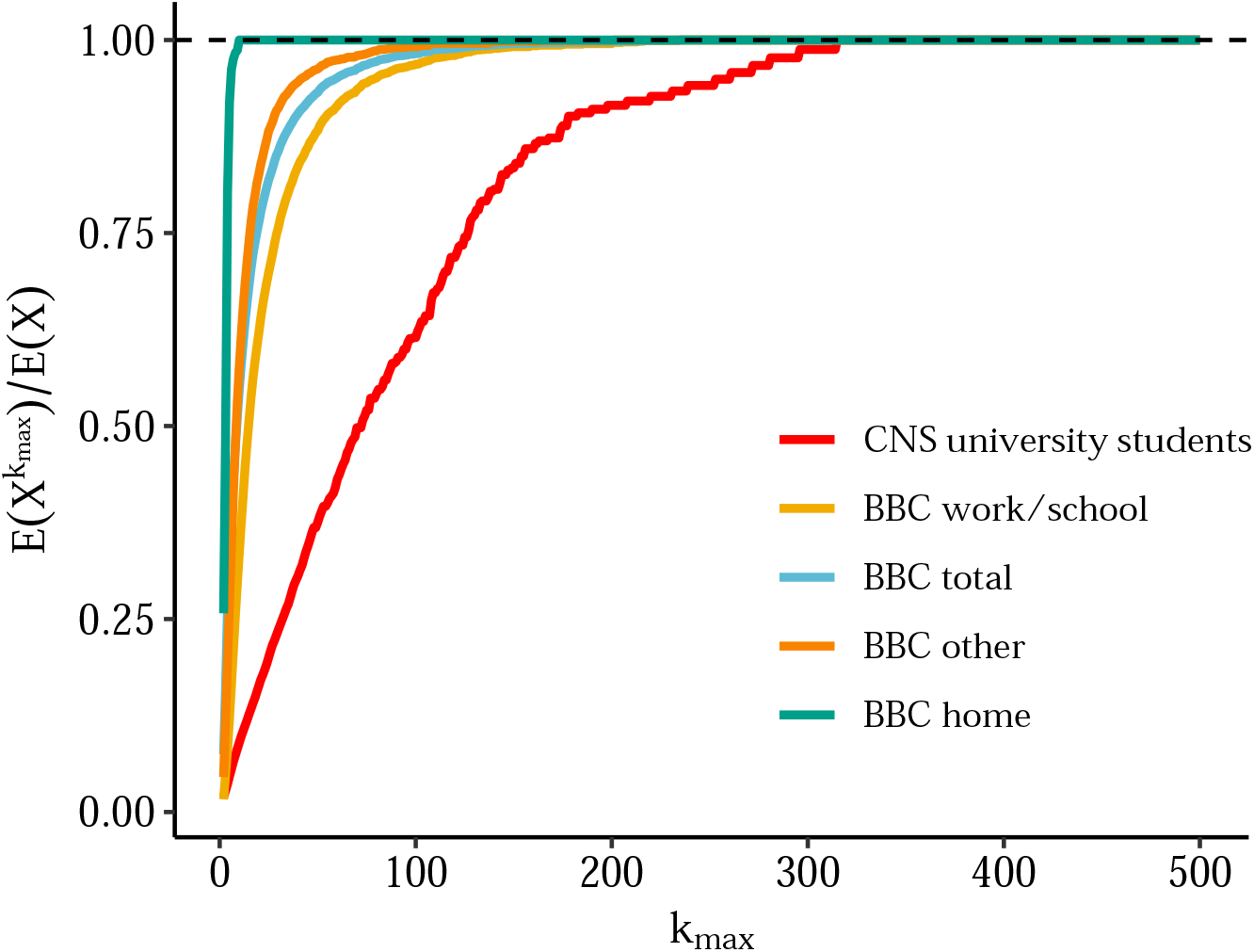
Relative rate of incident cases under restriction which prohibits gatherings above size *k*_*max*_ using different data sources for the distributions of gathering sizes. Again we fix transmission parameters to following values *p*_*i*_ = 0.01, *p*_*s*_ = 0.99, *τ* = 0.08 but use draws from empirical distributions in Figure 3. The relative rate of incident cases calculated using equation 6 and comparing restrictions with *k*_*max*_-level thresholds to unrestricted rate (e.g. a value of 0.5 implies a 50% fewer per capita incident cases at time *t* relative to unrestricted rate) is shown.

Finally, while taking the pre-restriction (and pre-outbreak) distribution such as in the analysis above can help one plan for extreme scenarios, it is clear that humans react to restrictions in complex ways that may not mirror the ideal discussed above. Therefore, we also extracted data from the CoMIX study (CMMID COVID-19 working group et al., 2020), which was designed as a deliberate follow on to the BBC Pandemic during the COVID-19 pandemic. In this study, a representative sample of 1,240 adults in the UK were asked about their contact patterns in the first week of the government-imposed ‘lockdown’ in March 2020. As before, we extracted the number of contacts made in a day by setting (home, work/school, or other). This additional data provides insight into the distribution of contacts under strong social distancing measures.

Figure 6 shows the full distribution of gathering sizes on a log-log scale comparing CoMIX to the pre-pandemic “normal” recorded in the BBC Pandemic study. Although sample sizes were considerably lower in CoMIX, several interesting patterns emerge. First, the distribution of household contacts under lockdown is almost identical to its pre-pandemic baseline, which is reassuring given household composition is largely unaffected by lockdown. Next, gatherings at work/school and other settings appear “clipped” relative to their pre-pandemic baseline and there now appears to be a preference for lower gathering sizes with a few outliers. This seems consistent with most people complying with order and a few who can’t (for instance because their occupation is among those deemed “essential”) or who refuse.

**Figure 6:**
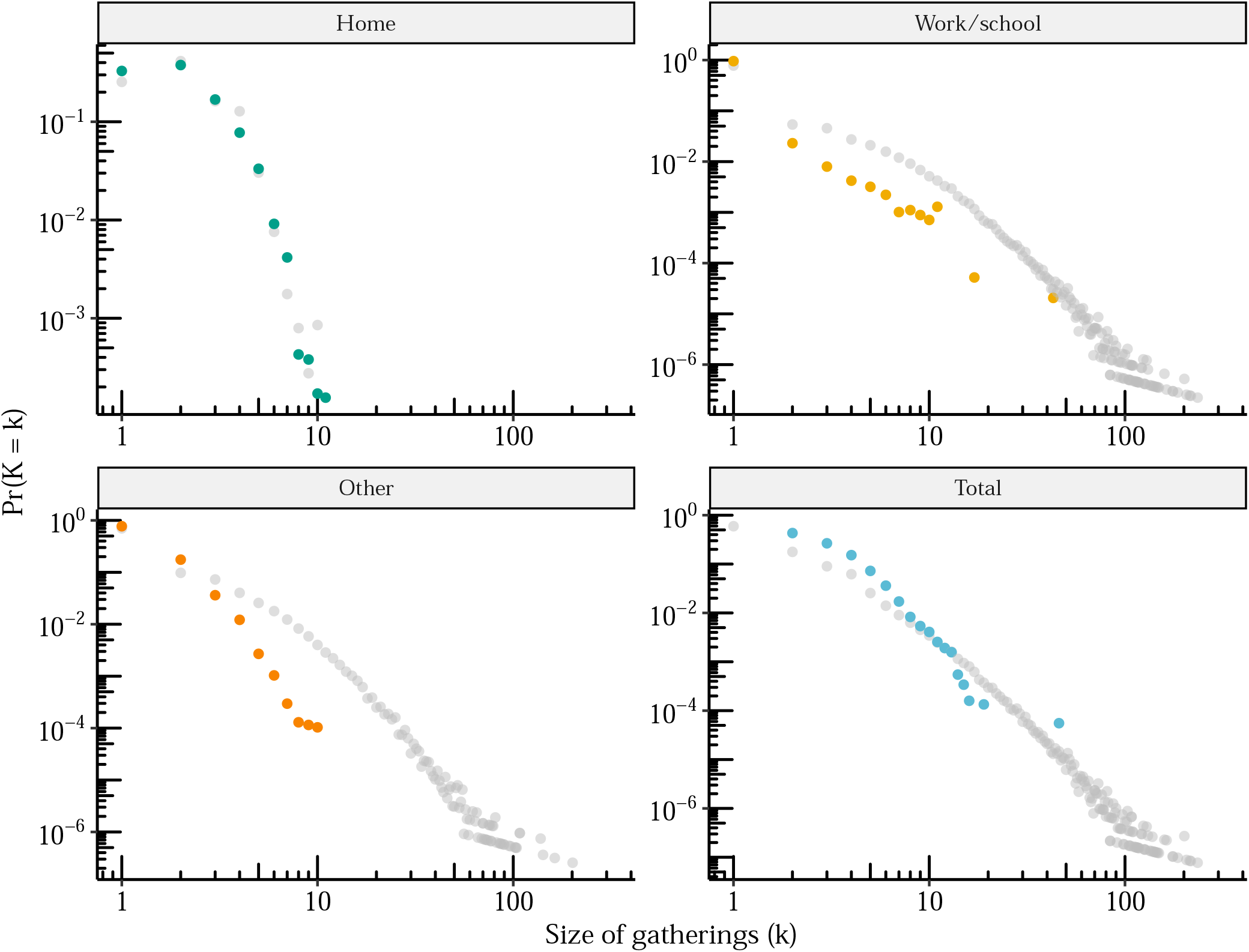
Distribution of gathering sizes before/during UK lockdown from the CoMIX study. Empirical distribution of gathering sizes by setting (home, work/school, other and total) during UK lockdown in March of 2020 are shown in color as measured in the CoMIX study. For comparison, the corresponding distributions as measured pre-pandemic in the original BBC Pandemic study are shown in gray for each setting.

## 4 Discussion

As the COVID-19 pandemic has demonstrated, non-pharmaceutical interventions are an essential tool to limit the spread of infectious diseases, both in the absence of vaccines or effective therapeutics, and when facing surges that test capacity of health systems or the emergence of new variants. We have shown that, when considering limitations on gathering size, decision-makers should consider the distribution of gathering sizes in addition to local conditions when determining the optimal threshold. While a lot of attention has focused on large gatherings, we show that small gatherings, due to their frequency, can be important contributors to transmission dynamics. Using empirical data from previous studies, we find that gathering size distributions are in fact “heavy-tailed” but that meaningful reduction in new cases only occurs once restrictions are set quite low. In theory this conclusion should also apply to future emerging variants of COVID-19 as well as future epidemics other than COVID-19. Our conclusion aligns with that of Brooks-Pollock et al. (Brooks-Pollock et al., 2021) who have showed that large gatherings of 50+ individuals have relatively small epidemiological impact while small and medium-sized groups of 10 to 50 individuals contribute most to COVID-19 epidemics.

Our work highlights the fact that more detailed data on human gathering sizes dynamics are needed, as datasets on this facet of social dynamics are extremely rare. This should include data on gathering size and duration across contexts and seasons as well as how distributions change during the course of an outbreak. These data, if available to policy-makers, would allow for more tailored restrictions and potentially more effective interventions. They would also contribute to better understanding of micro-dynamics of transmission during an outbreak and better parameterization of infectious disease models. Continuously tracked, remotely sensed data from cell phones, with appropriate anonymization and protection of individuals, may be one avenue for collecting this information on a large scale. Researchers and policy-makers could gain from increased access to such data.

Our model relies on multiple simplifying assumptions. Recognizing that violations are not equal and from the point of view of the policymaker the cautious approach is often the most prudent, where possible we have made effort to make conservative assumptions. First, by using a single probability we ignore many important heterogeneities in transmission risk (e.g indoor vs outdoor, use of face coverings, duration, ventilation, etc). However, as shown in A.2.1, this would only substantively affect our conclusions if heterogeneity varies with gathering size. For instance, if larger gatherings are more likely to be outdoor and people perceive them to be more dangerous and therefore adhere more strictly to masking and social distancing guidance then it’s possible that the per contact transmission risk may decrease with size of gathering, making restrictions on large gatherings even less effective relative to smaller ones. In this case, it would still be possible to apply the model presented, but specifying the transmission risk for each gathering size, which in practice may be hard to empirically validate.

Second, we assume in the main analysis that the probability of transmission is constant across gathering sizes, which may not be reasonable for very large gatherings (except perhaps in the case of an airborne pathogen in an unventilated and crowded indoor space). Here our model clearly represents a worst case scenario where all individuals have contacts with all other attendees. It thus likely overestimates the contribution of large gatherings to the overall number of new infections. We show in a sensitivity analysis in A.2.1 how allowing *τ* to vary with gathering size may affects our conclusions.

Third, we assume that susceptible, infected and recovered individuals are exchangeable, mix randomly, exhibit the same behaviour and attend gatherings at the same proportion as their proportion in the underlying population distribution. This may not be the case if, for instance, infectious people self-isolate upon developing symptoms or if there exists significant subsets of susceptibles who avoid gathering s and significant recovereds who believe they are immune and therefore go to gatherings at rates above their population fraction. Again these heterogeneities in behavior will mostly affect our conclusions if they vary with size of the gathering. In particular, they may lead to substantially different conclusions if behavioral dynamics tend to favor transmission at larger gatherings, such as if a infectious individuals are more likely to attend large gatherings. Even if they vary with gathering size though, they must also overcome the relative rarity of large gatherings. For instance, it is plausible that for a given “super-spreader”, i.e. an individual with enhanced infectiousness either due to biology, timing, or sociability, their impact would scale with gathering size. Indeed, some of the most well-publicized super-spreading events have occurred at large gatherings, such as choir practices (Hamner et al., 2020) or weddings (Mahale et al., 2020). However, these events are unlikely to contribute meaningfully to determination of restriction thresholds as they require the conjunction of two extremely rare events: a large gatherings occurring, and a very infectious ’super-spreader’ individual attending such a large gathering.

Similarly, in estimating effect of a certain threshold, we assume that individuals respond to gathering size restrictions uniformly, with perfect compliance and that they do not adapt their social behaviours independently of the regulation, based on, for instance, their knowledge of local epidemic dynamics. This is obviously not true in practice, but most plausible deviations would tend to make our estimates an upper bound on the effect of restrictions above a certain size. However, if announcing any restriction is a sufficient signal that many opt to avoid any gatherings at all, that may lead to a large reduction in cases even at a relatively large threshold. This may be more likely at the start of an outbreak when people are still attempting to ascertain the seriousness of the risk. In practice, responses to restrictions have varied, both across places and at different points during the pandemic as enthusiasm wanes (Kishore et al., 2022; Petherick et al., 2021). Policy-makers should take this into account when determining the right threshold.

Lastly, we assume all new infections to be equivalent, not considering heterogeneity in the impact of secondary infections. This assumption again may not be reasonable at the beginning of an epidemic when local transmission is not established and we might expect infections at larger gatherings to seed downstream cases in more diverse parts of the population/community. In this case although larger gatherings are less frequent they act as central nodes in the contact graph through which infection reaches sub-communities.

Our work is also subject to several limitations due to the data sources that we used. The three data sources (BBC Pandemic, CoMIX and CNS) had different aims, study designs and limitations. We assume they all provide good estimates of frequency of gathering sizes. In using the BBC Pandemic and CoMIX study we approximate the size of gatherings by assuming that all daily contacts in a given context all took place in one gathering. The CoMIX study was conducted during the first week of lockdown in March 2020 in the UK and may not be representative of restrictions in other locations or times. The Copenhagen Networks Study studied university students in Copenhagen, a specific population that may not be representative of other populations. In the case of the BBC Pandemic and CoMIX data, a particular threat to our main conclusions might be measurement error that correlates with gathering size, for instance if people get worse at recalling or recording the size of larger gatherings we may underestimate their frequency and therefore their contribution to transmission dynamics. This is a major advantage of the CNS data which were remotely recorded by cellphone and gps devices.

## Data Availability

All code and data produced are available online at : https://github.com/boyercb/covid-gathering-size

## A Appendix

### A.1 Theory

#### A.1.1 Derivation of relationship between expected cases and gathering size

Assuming fixed transmission probability *τ* for contacts between susceptibles and infectious individuals, the number of secondary cases generated by a single infectious individual with *K*_*s*_ = *k*_*s*_ susceptible contacts is binomially distributed, i.e.

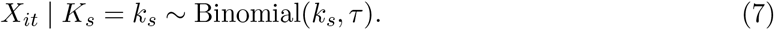

If susceptibles, infectious, and recovered individuals attend gatherings at rates equivalent to their population proportions then attendance at a gathering of size *K* = *k* can be represented by a multinomial sampling model of the form

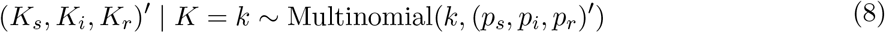

Where 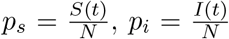, and 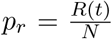. Under the model, the expected number of susceptibles is *kp*_*s*_, the expected number of infectious is *kp*_*i*_, and the expected number of recovereds is *kp*_*r*_. To calculate the expected number of secondary cases, note that, on average, only the *K*_*s*_ susceptibles are at risk of infection and they are exposed to *K*_*i*_ infectious individuals. Then the probability that the *K*_*s*_ susceptibles “escape”, i.e. that they are not infected by any of the *K*_*i*_ infectious individuals, is 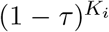 and thus, by extension, the probability that they are infected by at least one of the *K*_*i*_ infectious individuals in attendance is 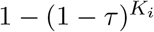. Therefore, the total number of secondary cases at a gathering of size *K* = *k* is

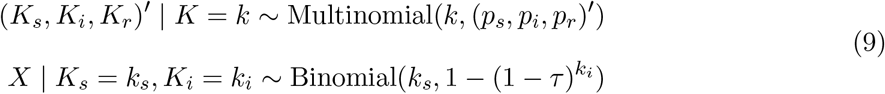

and taking iterated expectations, the expected number of secondary cases given a gathering of size *K* = *k* is simply

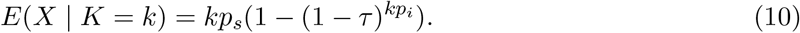

For a more intuitive way to think about this equation, notice that *kp*_*s*_ is the expected number of susceptibles and *kp*_*i*_ is the expected number of infectious individuals; the expression 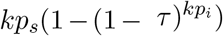 is then just the expected number of susceptibles times the probability of being infected by any of the infectious individuals who attend, where the latter is equivalent to the one minus the “escape” probability, i.e. the probability that no susceptible is infected by any of the infectious individuals expected to attend.

#### A.1.2 Binomial Approximation

More specifically, when |*τkp*_*i*_| ≪ 1 a Binomial approximation gives

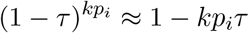

and thus

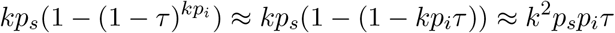

### A.2 Sensitivity analyses

In the main analysis, for simplicity of presentation and to fix concepts, we consider fixed values of *τ* as well as *p*_*s*_, *p*_*i*_, and *p*_*r*_. While we attempted to use values consistent with the literature for COVID-19, in practice these may vary across settings and for different pathogens. Here we conduct a series of sensitivity analyses to explore the role that these parameters play in determining impact of gathering size restrictions. In the main text, we assumed that the distribution of gatherings follows a discrete power law distribution. Here, we instead consider another heavy-tailed distribution, the log-normal distribution and show it fits to the various empirical data.

#### A.2.1 Varying *τ*

The probability of transmission given contact, *τ*, can vary either in constant value —for instance, if a new variant emerges that is more infectious— or more often it may simply be heterogeneous across settings —for example, a crowded indoor gathering versus an outdoor gathering. In the case of the former, in Figure A.1 we range *τ* over a suitable range, for instance 0.01 to 0.25 and find that our results are not substantially changed. We chose 0.01 as a lower bound, 0.08 as a medium value from the secondary attack rate during meals (Bi et al., 2020), and 0.25 as a higher bound from the secondary attack rate in households during the Omicron wave (Jørgensen et al., 2022). In the case of the latter, we can consider *τ* to be drawn from a distribution reflecting the population of gathering settings at any given time. Given that *τ* must be between 0 and 1, a natural starting point for incorporating heterogeneity in *τ* into our prior model is to draw it from a beta distribution, i.e.

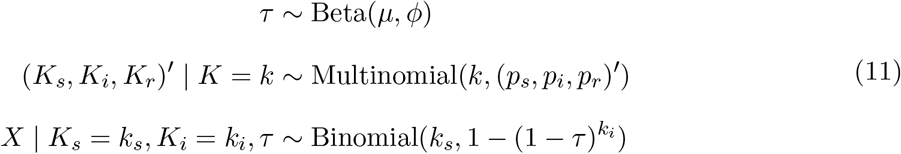

where here we’ve parameterized it such that *µ* is the mean of the beta distribution and *ϕ* is a dispersion parameter representing how concentrated values of *τ* are around the mean^1^. For now, we assume that *τ* is independent of gathering size. Figure A.2 shows example draws from beta distributions with same value of *τ* but different dispersion. As the dispersion parameter increases, *τ* is increasingly concentrated around the mean, as it decreases *τ* is more variable. In practice, increasing variability in *τ* would indicate that there are a small number of gatherings with very high transmission and a larger number with little to no transmission.

We show the effects of dispersion in *τ* on our conclusions regarding gathering size restrictions in Figure A.3. Given that our derivation of equation 10 relies only on the mean of *τ* and we assume that tau is independent of gathering size, we should expect that the expected reduction in incident cases for a *k*_*max*_ restriction is unchanged by varying *ϕ* as

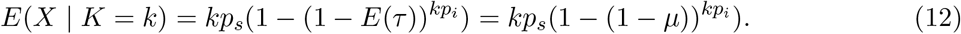

Practically, this implies that as long as *τ* is independent of gathering size (and the mean value of *τ* is defined) our long run conclusions about the effect of gathering size restrictions is unchanged. However, as highlighted by the shaded regions of the 95% simulation intervals, increasing variability in *τ* leads to greater variance in the effect of restrictions. In terms of policy-making, if there’s strong evidence for heterogeneity in *τ* decision-makers may want to consider planning with these intervals in mind (e.g. using upper bound from an appropriately chosen interval such that there’s an *p*% chance that restriction leads to reduction of desired size).

**Figure A.1:**
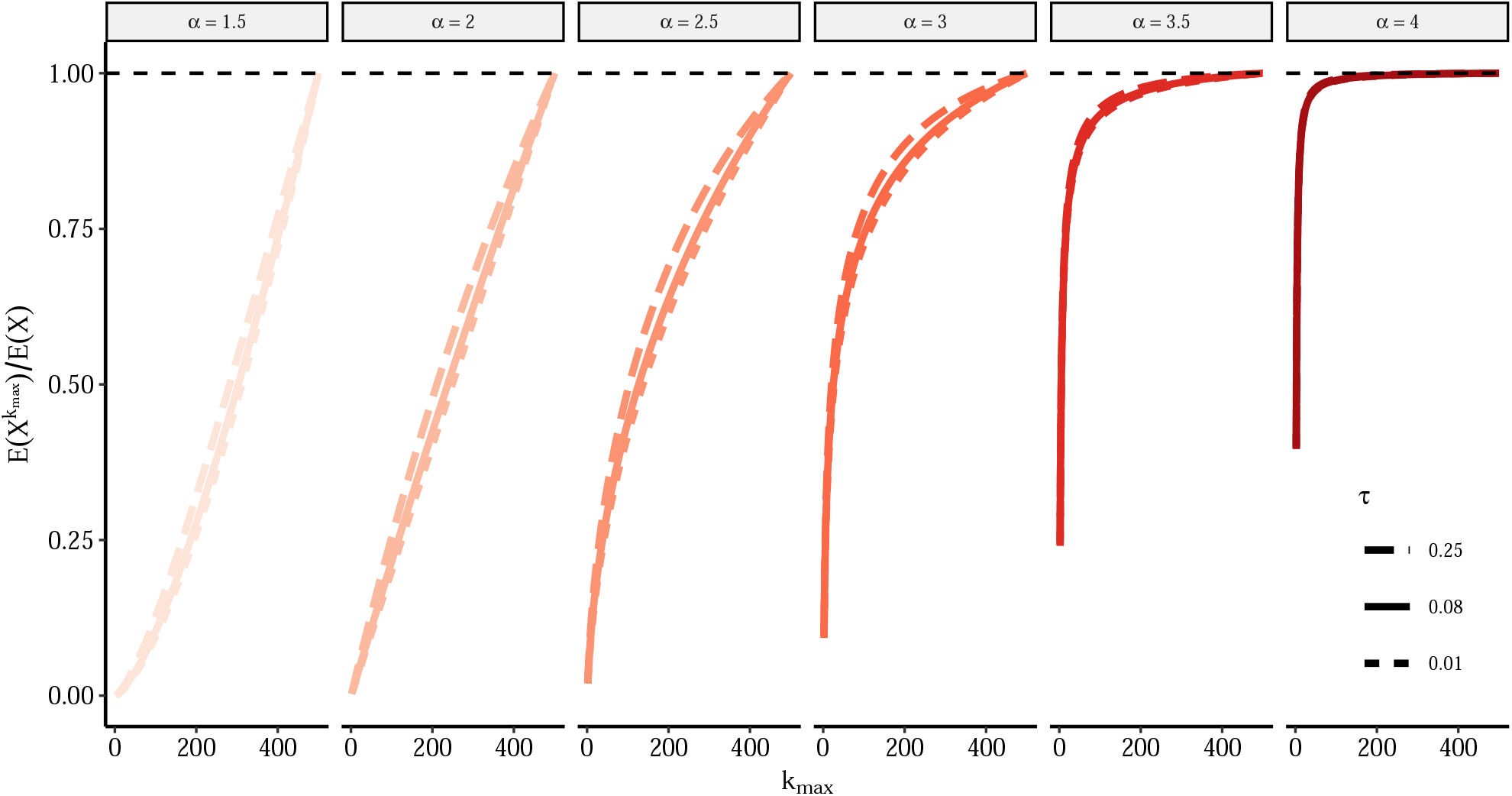
Effect of different values of constant *τ* on relative rate of incident cases under *k*_*max*_ gathering size restrictions for different power law distributions. Here we re-create Figure 2 but vary the values of the constant *τ* from 0.01 to 0.25.

**Figure A.2:**
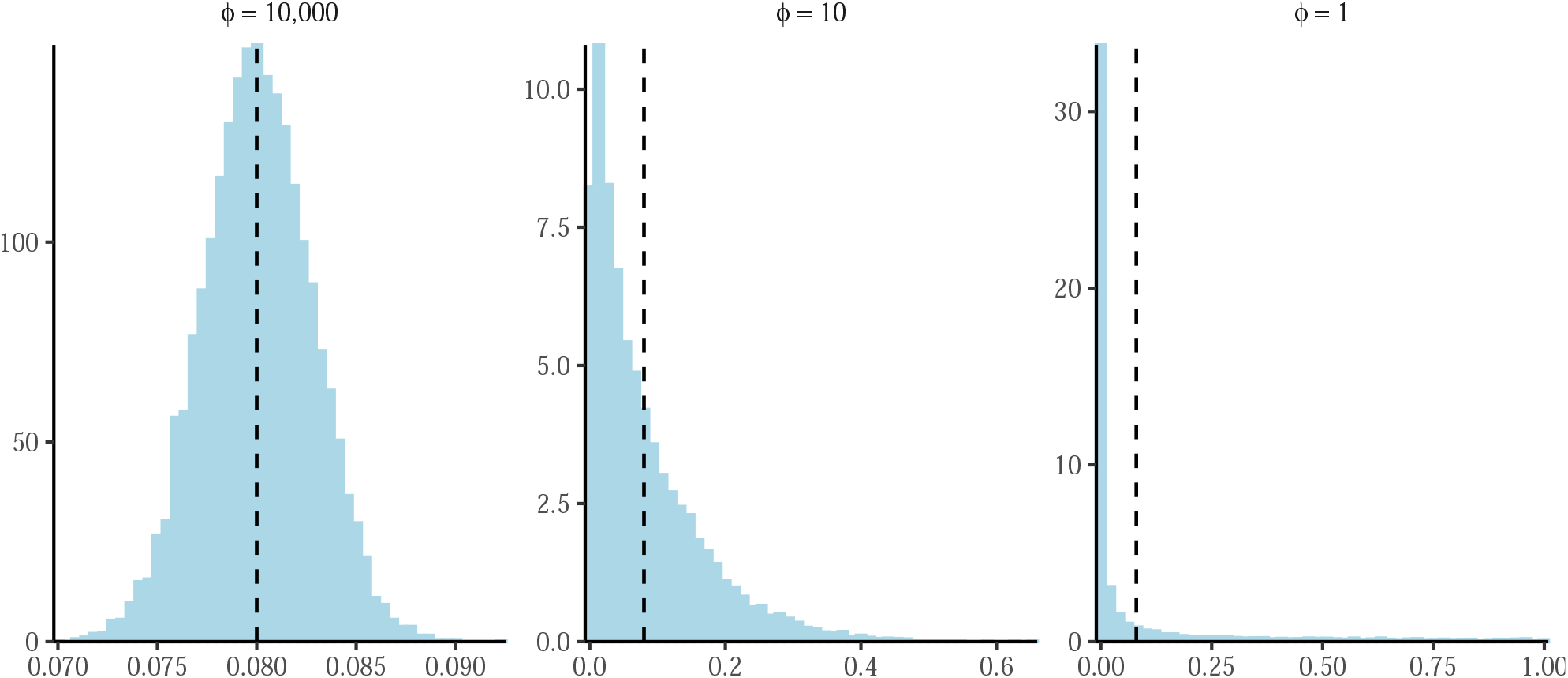
Examples of the beta distribution with *µ* = 0.08 under different values of *ϕ*. Based on 10,000 draws from the beta distribution with *µ* fixed to 0.08; dashed line shows the position of *µ*.

**Figure A.3:**
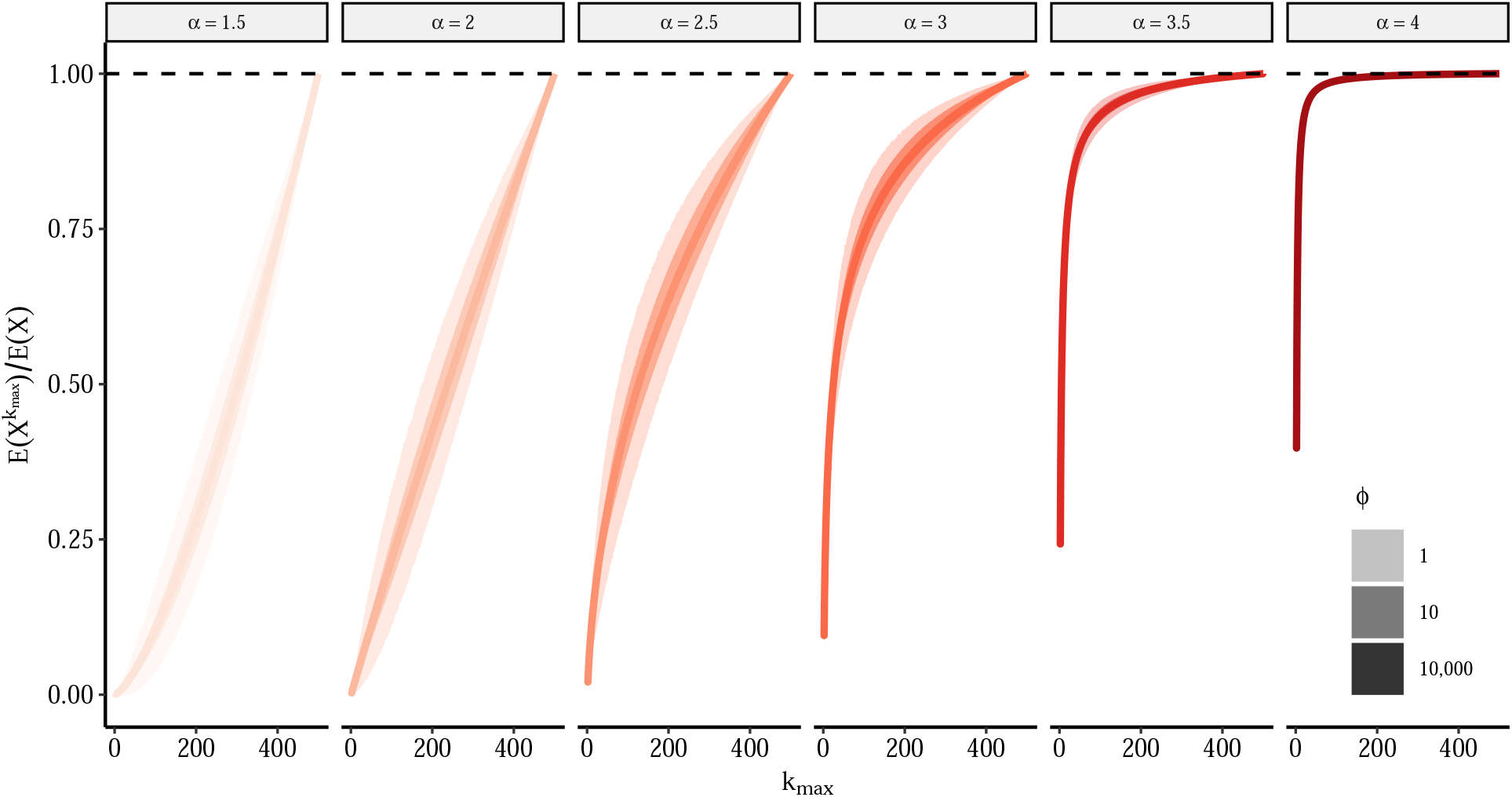
Effect of dispersion in *τ* on relative rate of incident cases under *k*_*max*_ gathering size restrictions for different power law distributions. Here we fix transmission parameters to following values *p*_*i*_ = 0.01, *p*_*s*_ = 0.99, but allow *τ* to vary. We draw 10,000 values of *τ* from a beta distribution with *µ* = 0.08 and varying *ϕ*. As previously, the figure shows the relative rate of incident cases calculated using equation 6 and comparing restrictions with *k*_*max*_- level thresholds to unrestricted rate (e.g. a value of 0.5 implies a 50% fewer per capita incident cases relative to unrestricted rate). The lines represent the mean relative rate across all simulations while the shaded areas show the 95% simulation intervals.

Finally, we consider what happens when *τ* is allowed to vary with the size of gathering. Given the paucity of data, it’s unclear what one might expect the relationship to be between *τ* and gathering size *a priori*. On the one hand, *τ* could increase with gathering size if larger gatherings tend to be longer or in settings more conducive to spread. On the other hand, one could make an equally compelling case that smaller gatherings, which may be in more intimate settings may have higher *τ*. In their paper describing the Copenhange Network Study data, (Sekara et al., 2016) find no association between the size and duration of gatherings, suggesting no relationship on at least one proxy for *τ*. Absent reliable sources, here we vary the relationship across three representative scenarios: (1) *τ* decreases with gathering size *K*, (2) *τ* independent of gathering size *K, τ* increases with gathering size *K*. To keep it simple, we group gatherings into three mostly arbitrary groupings (less than 10 people, 11 to 50 people, and larger then 51 people) and choose *τ* for each from *τ* ∈ {0.01, 0.08, 0.25}. Figure A.4 shows the results. Relative to our results presented in the main text, represented by the solid line where *τ* is independent of *k*, if *τ* decreases with gathering size our estimates are too optimistic and harsher restrictions would be required to achieve the same reductions. If *τ* increases with gathering size, then our estimates are too pessimistic and comparable reductions could be achieved with looser restrictions on size.

**Figure A.4:**
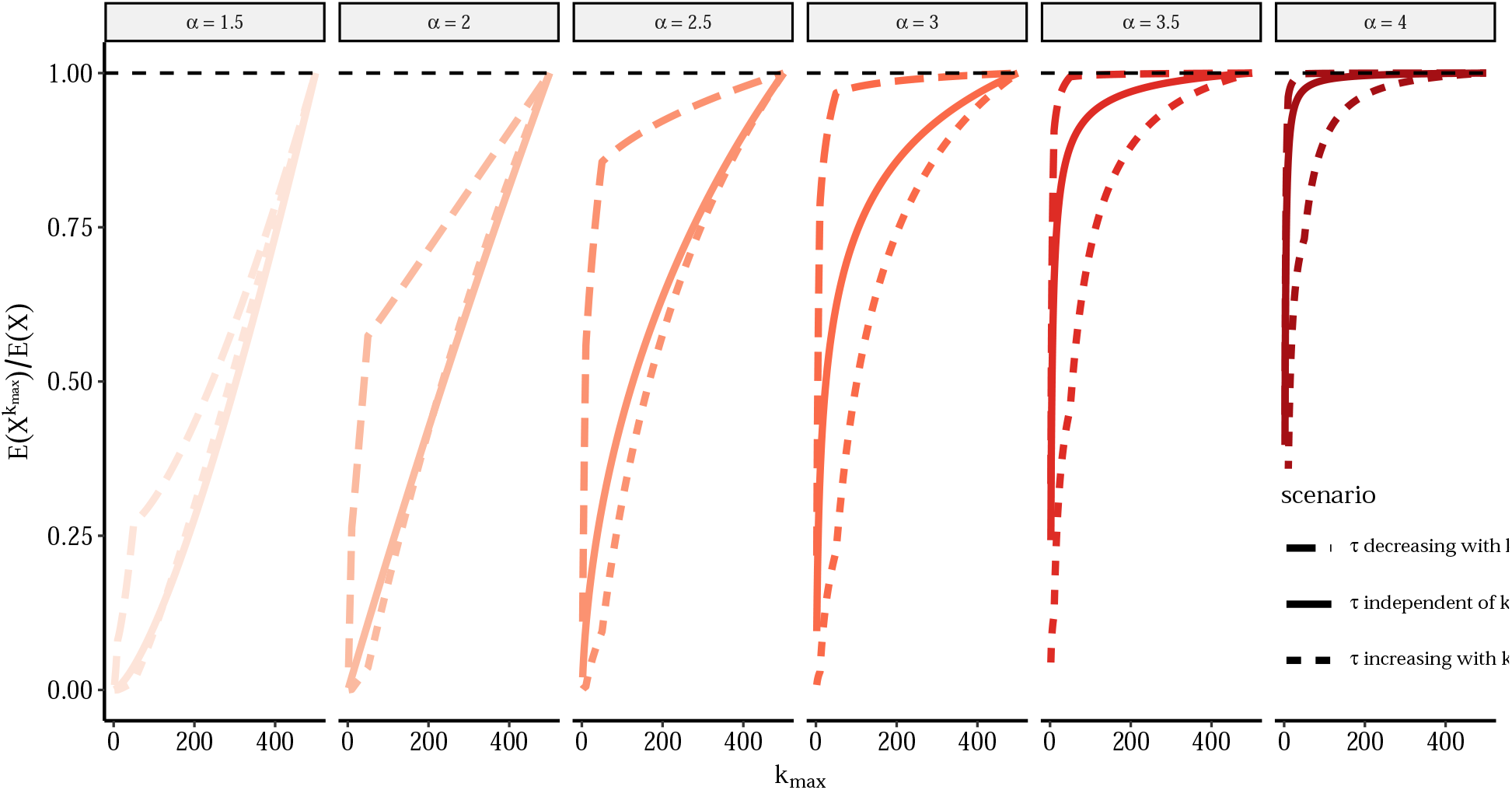
Effect of relationship between *τ* and gathering size *k* on relative rate of incident cases under *k*_*max*_ gathering size restrictions for different power law distributions. Here we consider three scenarios for how the transmission probability *τ* varies with gathering size *k*. For *τ* decreasing with *k, τ* = 0.25 for gatherings with less than 10 people, *τ* = 0.08 for gatherings with 11 to 50 people, and *τ* = 0.01 for gatherings with more than 51 people. For *τ* increasing with *k, τ* = 0.01 for gatherings with less than 10 people, *τ* = 0.08 for gatherings with 11 to 50 people, and *τ* = 0.25 for gatherings with more than 51 people. The solid line is the reference case where *τ* = 0.08 at all sizes.

#### A.2.2 Varying *p*_*i*_ and *p*_*r*_

The proportion of susceptible, infected and recovered individuals in each gathering are inputs to our gathering size restriction equation. In the main analysis, we fix values of *p*_*s*_, *p*_*i*_ and *p*_*r*_ to 0.99, 0.01 and 0 respectively, which seemed reasonably illustrative for our purposes. However, in sensitivity analyses presented below, we vary the values of *p*_*i*_ between 0.001, 0.01 and 0.1; and that of *p*_*r*_ between 0, 0.25 and 0.75. Across all nine combinations of *p*_*i*_ and *p*_*r*_, the value of *p*_*s*_ is equal to 1 − (*p*_*i*_ + *p*_*r*_) and thus varies from 0.15 to 0.999.

As shown in Figure A.5, for different power law distributions, the value of *p*_*r*_ seems to have only a small impact on the relative rate of incident infections comparing a scenario with restrictions to scenarios without. However, values of *p*_*i*_ seem to have a larger effect, especially for power law distributions with smaller *α* values. For very high proportions in infected individuals (i.e. *p*_*i*_ = 0.1), a given gathering size restriction leads to a lower relative reduction in the number of incident cases, when compared to lower proportion on infected individuals (i.e. *p*_*i*_ = 0.01 or 0.001). This makes sense as with such high proportion of infected attendees, and under distributions of gathering sizes where larger gatherings are frequent, most large gatherings will lead to a substantial number of infections. It is only when considering distribution of gatherings where larger gatherings are rare (i.e. *α* = 3.5 or 4) that the proportion of infected individual attending *p*_*i*_ matters less, as in smaller gatherings less individuals can be newly infected.

As shown in Figure A.6, across all five empirical distribution of gathering sizes, the values of *p*_*i*_ and *p*_*r*_ have very limited impact on the the relative rate of incident cases under restriction to gatherings of size *k*_*max*_ compared to that in absence of restrictions. Indeed, the dashed lines representing the varying values of *p*_*i*_ overlap in most plots, only showing a slight shift when using the distribution derived from CNS data source. Similarly, ranging values of *p*_*r*_ also appears to have minor impact.

Overall, these results suggest that the proportion of susceptible, infected and recovered individuals attending gatherings has limited to no effect on our relative reduction in incident cases when implementing restrictions. This suggests that our results may be robust to those parameters that our conclusion may hold for various stages of epidemics.

#### A.2.3 Alternative gathering size distributions

In the main text we illustrate our theoretical points using the discrete power law distribution and then follow them up with results using empirical distributions. In practice we find that, while informative from a theoretical standpoint, a power law may not provide the best fit empirically. Therefore, when informing policy, rather than intuition, we encourage the use of realistic and preferably empirically-derived distributions. However, such data may not always be available and thus for completeness here we also consider other heavy-tailed distributions. A relatively common heavy-tailed alternative is the log-normal distribution, i.e.

**Figure A.5:**
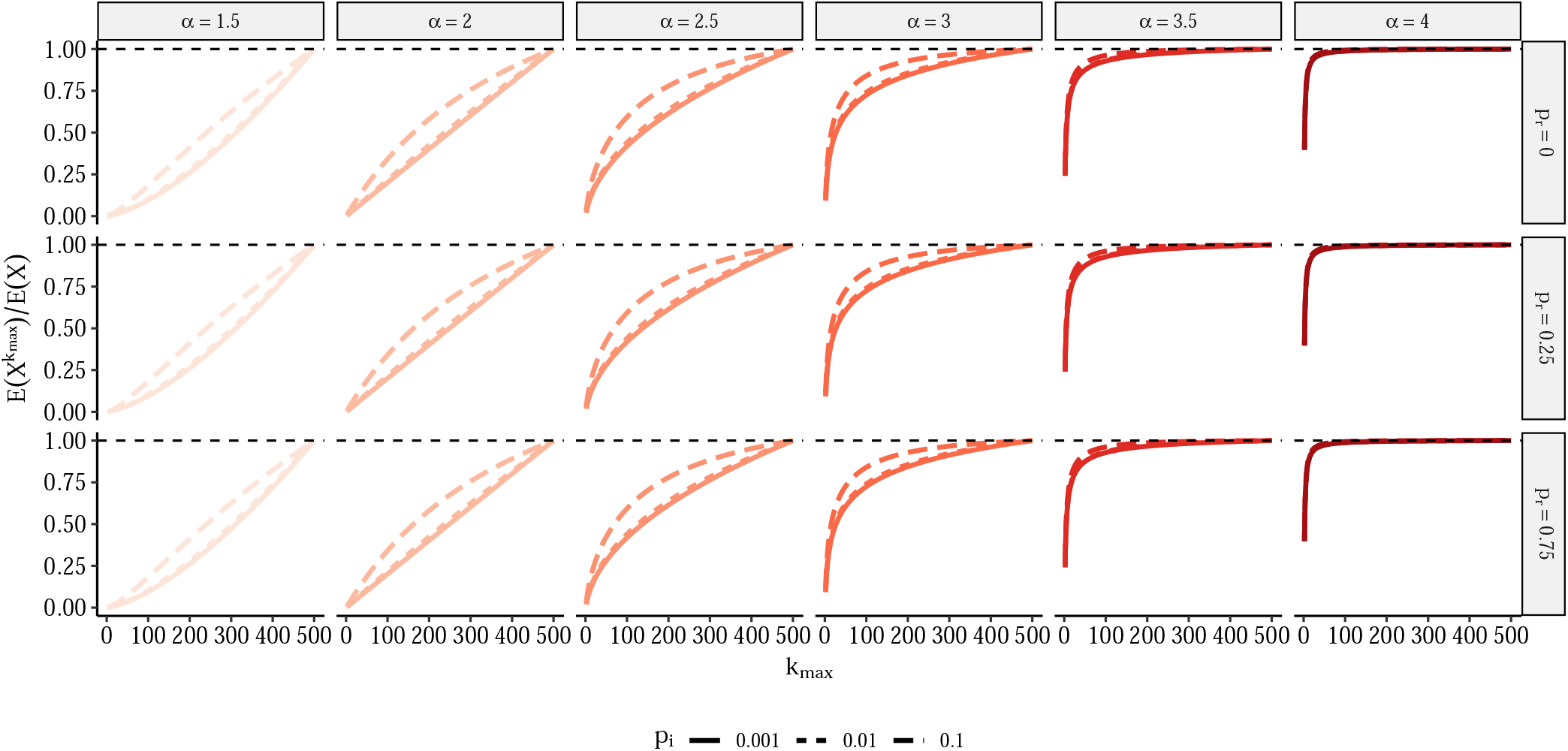
Relative rate of incident cases under restriction which prohibits gatherings above size *k*_*max*_ for different power law distributions and varying the values of *p*_*i*_ and *p*_*r*_. The value of *p*_*i*_ varies between 0.001, 0.01 and 0.1. That of *p*_*r*_ varies between 0, 0.25 and 0.75 and *p*_*s*_ is equal to 1 − (*p*_*i*_ + *p*_*r*_). *τ* is equal to 0.08. Similarly to Figure 2, we assume that power law behavior starts at *k*_*min*_ = 1 and truncate the power law above gatherings of size 500 both to make the sum tractable and given that gathering sizes must at minimum be less than population size. The figure shows the relative rate of incident cases calculated using equation 6 and comparing restrictions with *k*_*max*_-level thresholds to unrestricted rate (e.g. a value of 0.5 implies a 50% fewer per capita incident cases at time *t* relative to unrestricted rate).

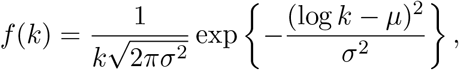

where the parameters *µ* and *σ*^2^ describe the mean and the variance of a log-transformed normal random variable. Figure A.7 provides some examples of parameter values with increasing tail mass.

**Figure A.6:**
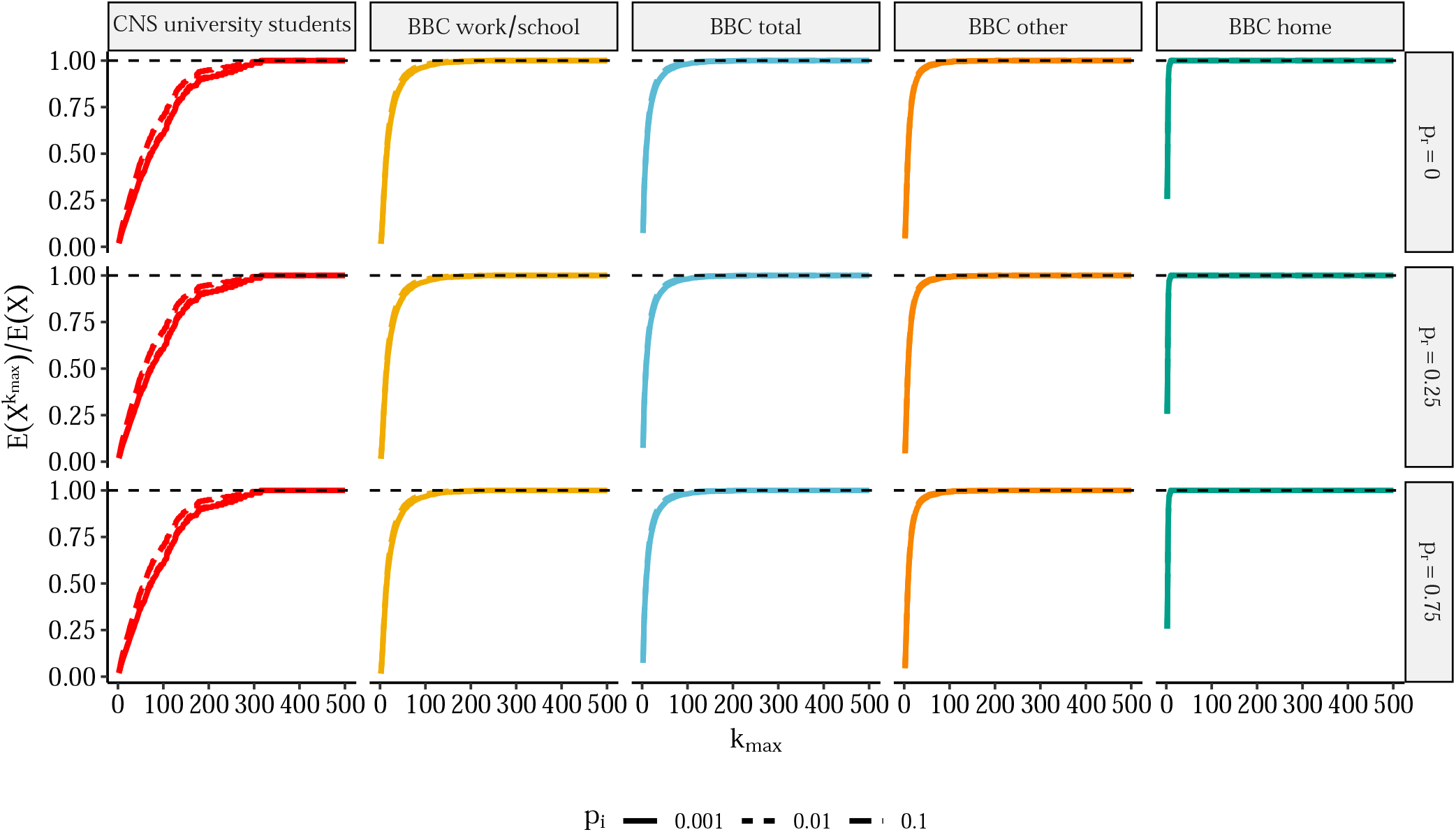
Relative rate of incident cases under restriction which prohibits gatherings above size *k*_*max*_ using different data sources for the distributions of gathering sizes and varying the values of *p*_*i*_ and *p*_*r*_. The value of *p*_*i*_ varies between 0.001, 0.01 and 0.1. That of *p*_*r*_ varies between 0, 0.25 and 0.75 and *p*_*s*_ is equal to 1 − (*p*_*i*_ + *p*_*r*_). *τ* is equal to 0.08. Similarly to Figure 5, we use draws from empirical distributions. The figure shows the relative rate of incident cases calculated using equation 6 and comparing restrictions with *k*_*max*_-level thresholds to unrestricted rate (e.g. a value of 0.5 implies a 50% fewer per capita incident cases at time *t* relative to unrestricted rate).

**Figure A.7:**
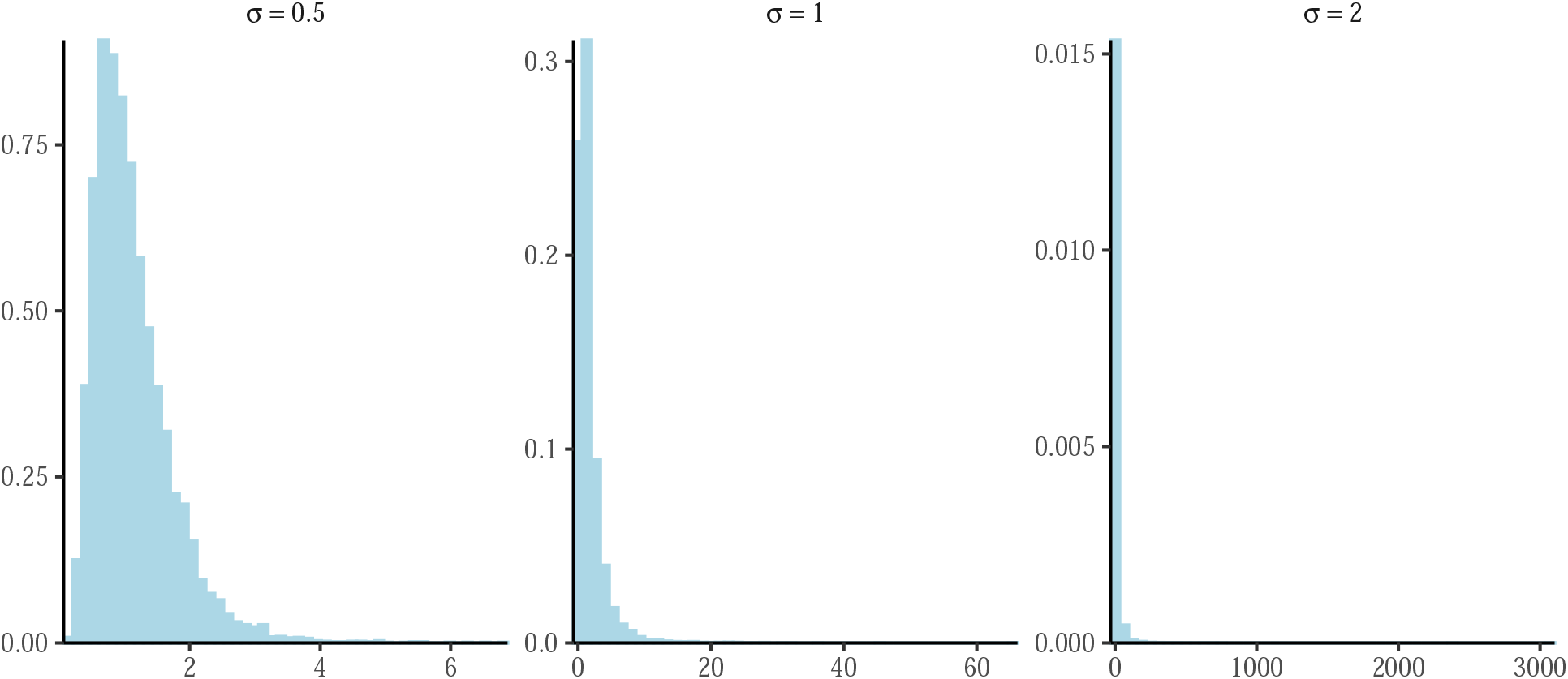
Examples of the log-normal distribution with *µ* = 0 under different values of *σ*. Based on 10,000 draws from the beta distribution with *µ* fixed to 0.

As in the main text, we fit the log-normal distribution to the empirical distributions from the BBC Pandemic and Copenhagen Networks Study using maximum likelihood. We again consider the possibility that the empirical data may only follow a log-normal distribution above a certain lower threshold (*k*_*min*_). Figure A.8 shows the best fitting log-normal distributions and their parameter values. Visually the log-normal distribution seems like a better fit than the discrete power law considered in the main text, mostly because there does seem to be some nonlinear drop-off in the extreme tails. However, this could also reflect influence of measurement error^2^ and sampling variability in the extreme tail. We can test this using Vuong’s likelihood ratio test which compares the Kullback-Leibler criteria for the two fits. In Table A.1, we find strong evidence that the log-normal is a better fit for all but the household contacts.

**Figure A.8:**
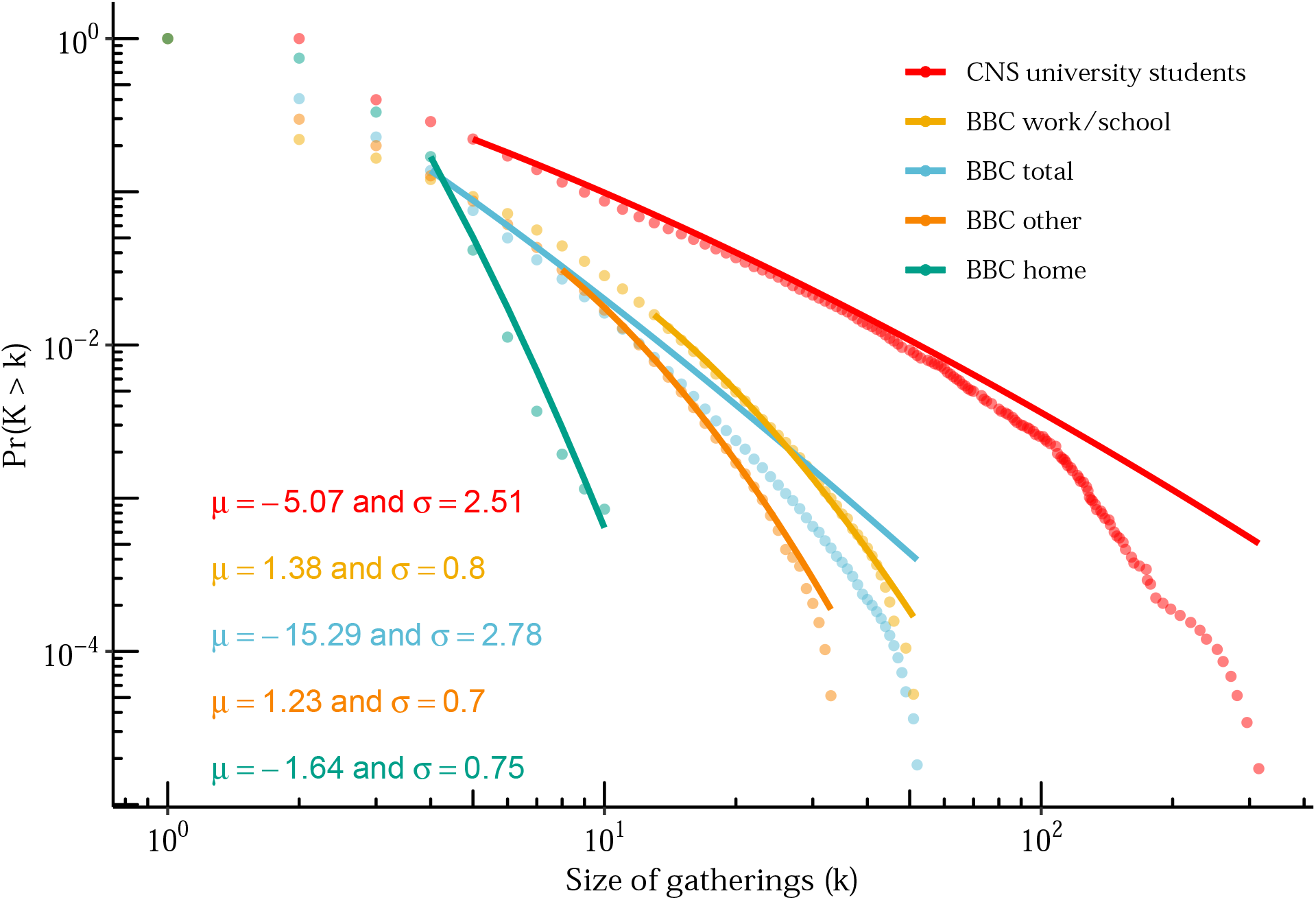
Estimates of log-normal parameters for the Copenhagen Networks Study (CNS) and the BBC Pandemic study by setting. Plot is complementary cumulative distribution function versus gathering size with lines showing fitted power law distribution. Estimates for *α* and *k*_*min*_ obtained using maximum likelihood for discrete power law using the `poweRlaw` package in R.

**Table A.1:** Vuong’s test comparing discrete log-normal and power-law distribution fits to empirical data on gathering size.

| Data source | $R$-statistic | $p$-value |
| --- | --- | --- |
| BBC Pandemic |  |  |
| Home | 1.24 | 0.216 |
| Work / school | 6.66 | <0.001 |
| Other | 5.37 | <0.001 |
| Total | 4.20 | <0.001 |
| Copenhagen Networks Study | 5.89 | <0.001 |

*Notes:* $R$ -statistic is the ratio of log-likelihoods of the discrete log-normal and power-law fits, positive values favor the log-normal distribution. $p$ -values are for two-sided hypothesis that log-normal is a better fit.

## Footnotes

1 The canonical definition of the beta distribution is in terms of shape parameters *α* and *β* where 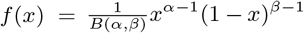. Here we use the following transformation 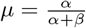 and *ϕ* = *α* + *β*, where *ϕ* is sometimes also called the sample size.

2 For instance, BBC Pandemic dataset has individuals record their daily contacts. It’s likely that they are better at estimating small group sizes relative to big ones and may lump or round estimates for larger groups.

